# Parental administration of analgesia to children attending the emergency department with acutely painful conditions

**DOI:** 10.1101/2020.12.15.20248274

**Authors:** T. Daly, R. O’Brien, A. Murphy

## Abstract

**Background:** Acute paediatric pain management is often suboptimal in the emergency setting. There is a paucity of published literature on parental administration of analgesia to children, prior to their arrival at the emergency department (ED). The objective of this study was to describe the prevalence of pre-hospital analgesic administration by parents/guardians of children ≤16 years presenting to ED, with acutely painful conditions.

**Methods:** This was a prospective cross-sectional study conducted across two ED’s in the South of Ireland with a collective annual attendance of approximately 100,000 patients. A consecutive sample of 400 parents/guardians of children aged between 6 months and 16 years, who were self-referred to ED with acutely painful conditions, were included. Data collected included patient demographics, pain score and analgesia administration. Data was analysed with SPSS software using descriptive and inferential statistics.

**Results:** 189/400(47%) children received analgesia from their parents/guardians pre-ED arrival. Factors independently associated with increased parental administration of analgesia were: pain score ≥5/10 53.4%(95%CI 48%-59%) vs ≤4/10 29%(95%CI 21%-38%), children with siblings 49.7%(95%CI 45%-56%) vs without siblings 35.3%(95%CI 24%-47%) and presenting <48 hours from onset of pain 50.1%(95%CI 45%-55%) vs presenting ≥48 hours 30.5%(95%CI 19%-42%). Of the 400 participants, 211(53%) received no analgesia before attending ED. Reasons for parents not administering analgesia included: 62/211(29.4%) did not think the child needed it, 39/211(18.5%) accident did not happen at home, 34/211(16.1%) did not want to mask the presence of the pain, 20/211(9.5%) believed the hospital should give medications, and 18/211(8.5%) afraid it would be wrong/harmful.

**Conclusions:** Over half the children presenting to ED, with acutely painful conditions, did not receive adequate or timely pain relief pre-ED arrival, causing avoidable suffering. Parental misconceptions about acute pain management are major barrier to analgesic administration. Educational strategies are required to dispel misconceptions, which may ultimately improve the care for this population.

## Introduction

The World Health Organisation (WHO) has long declared adequate pain management a human right.^1^ Pain is the most common symptom in the emergency setting, with up to 70% of patients attending the Emergency Department (ED) having pain as part of their primary complaint.^2^ The international literature indicates that up to 64.1% of ED paediatric patients suffer pain as their chief complaint.^3^ However, the same study found that only 15% of those who reported pain received sufficient and timely analgesia. Numerous studies have highlighted the perceived inadequacies with acute pain management in paediatric patients attending the ED.^4, 5, 6^

The term “oligoanalgesia” was first used in 1989 by Wilson and Pendleton to describe the underuse of pain medication in the emergency department.^7^ Oligoanalgesia is known to have negative physical, biochemical, and psychological effects, sometimes with enduring consequences for paediatric patients and can negatively impact their future experiences of pain.^8^ Associations between early experiences of pain and numerous negative behavioural and psychological consequences later in life have been identified.^9^ Prior studies show that early experiences of pain can negatively affect future responses to pain into adulthood while appropriate pain control shows reduced changes in future pain behaviours.^10^

In spite of these long-established consequences, paediatric pain is consistently less recognised, documented and treated compared to adult populations.^6, 11^ Even in cases where acute pain is assessed, it is frequently underestimated and insufficient administration of pain relief to children has been observed in emergency medical services (EMS) providers, physicians and parents.^3, 12, 13^ There is a clear paucity in the literature on parental pre-hospital analgesia administration, with the majority of the literature focusing on the practices of EMS. This is despite a large proportion of children being self-referred to the ED with parents/guardians as the only pre-hospital carer providers. Acute pain management in children has not commanded the same level of research interest in contrast to the adult population, and recent studies have prioritised the need for high-level evidence and further research.^14^ Understanding current parental practices pertaining to paediatric pre-hospital analgesia may highlight gaps in knowledge and illuminate how it can be improved in the future.

## Objectives

The primary outcome of this study is to determine the prevalence of parent/guardian administration of analgesic medication to their children for the treatment of acutely painful conditions (i.e. trauma and non-trauma causes) prior to emergency department arrival. We also sought to determine analgesia types & doses, if administered. Reasons for not administering analgesia were also explored. Lastly, we examined associations between socio-demographic factors and the likelihood of parents/guardians administering pre-hospital analgesia.

## Methods

### Study Design and Setting

A prospective cross-sectional study was undertaken of paediatric patients who attended two EDs in the south of Ireland (Cork University Hospital (CUH) and Mercy University Hospital (MUH), Cork), Ireland between May and September 2019. CUH and MUH serves a catchment population of over 1.1 million. Collectively, over 100,000 patients seek care at CUH and MUH EDs annually. Ethical approval was sought and granted by the Clinical Research and Ethics Committee (CREC) for Cork Teaching Hospitals in February 2019.

### Selection of Participants

Systemic sampling was used to recruit a defined proportion of consecutively approached parents of children who met the eligibility criteria. To be eligible participants had to be parents/guardians of children aged 6 months – 16 years, with an acutely painful condition, who self-referred to the ED.

### Data Collection

The questionnaire employed is based on one used by a similar study; ‘Who gives pain relief to children’ performed in 1998 by Spedding et al.^15^ It was adapted to better suit the research question. After the questionnaire was modified, a pilot study was conducted to ensure the measurement instrument was suitable. The following data points were collected; Patient demographics, number of siblings, cause of pain (i.e. injury/illness), site of pain, pain score on ED arrival, whether analgesia was administered prior to ED arrival, if yes, what type and dose of analgesia administered, if no, reasons why analgesia was not administered.

### Study Procedure

Parents of children who met the eligibility criteria were approached in the ED waiting area following triage while waiting to be seen by an ED doctor. The parents were given a study invitation letter and a patient information letter to read which explained the purpose of the study and their role in it. They were asked to verbally consent to participation in the study. Participants then completed an anonymous questionnaire using an electronic tablet provided by the research team using Survey Monkey software. One questionnaire was distributed per child.

### Data Analysis

Data was exported to Microsoft Excel, and subsequently transferred to SPSS (Statistical Product and Service Solutions Version 26.0, SPSS Inc., Chicago, Illinois, USA) for analysis. Simple descriptive statistics were used to describe the clinical characteristics of the study population. Primary statistical analysis included assessment of the rate of pre-hospital analgesia administration, types and doses of analgesics administered and reasons for analgesia not being administered using summary statistics. Secondary statistical analysis involved examining associations between socio-demographic factors and the likelihood of parents/guardians to administer pre-hospital analgesia, using simple descriptive statistics, chi-squared test and odds ratios. A p-value of <0.05 was considered significant.

The last study of this nature performed in Ireland documented a prevalence of 24%. The sample size for this study was determined using a precision-based sample size calculation assuming an incidence of 25% for parent administration of pre-hospital analgesia. It was estimated 288 participants would be needed. At the mid-point in the study the incidence of parental administration of analgesia was higher than expected (approximately 50%) and so the sample size calculation was performed again with a new estimate of 384 participants required to be 95% confident that the percentage of parents administering pre-hospital analgesia was within +/-5% of the estimated value.

### Missing Data

Each of the 400 surveys were administered by one data collector who was present for each survey to support the parent/guardian. The survey was completed using Survey Monkey software on a tablet provided by the research team and took on average 5 minutes and 17 seconds to complete. Each survey was reviewed immediately by the data collector and additional information was sought for incomplete sections. As survey monkey does not allow submission of a survey unless all fields are filled there was very little missing data. There were 15 surveys with either missing data, surveys incorrectly filled out or participants were not eligible for the study and should not have been recruited. These responses were removed from the data set.

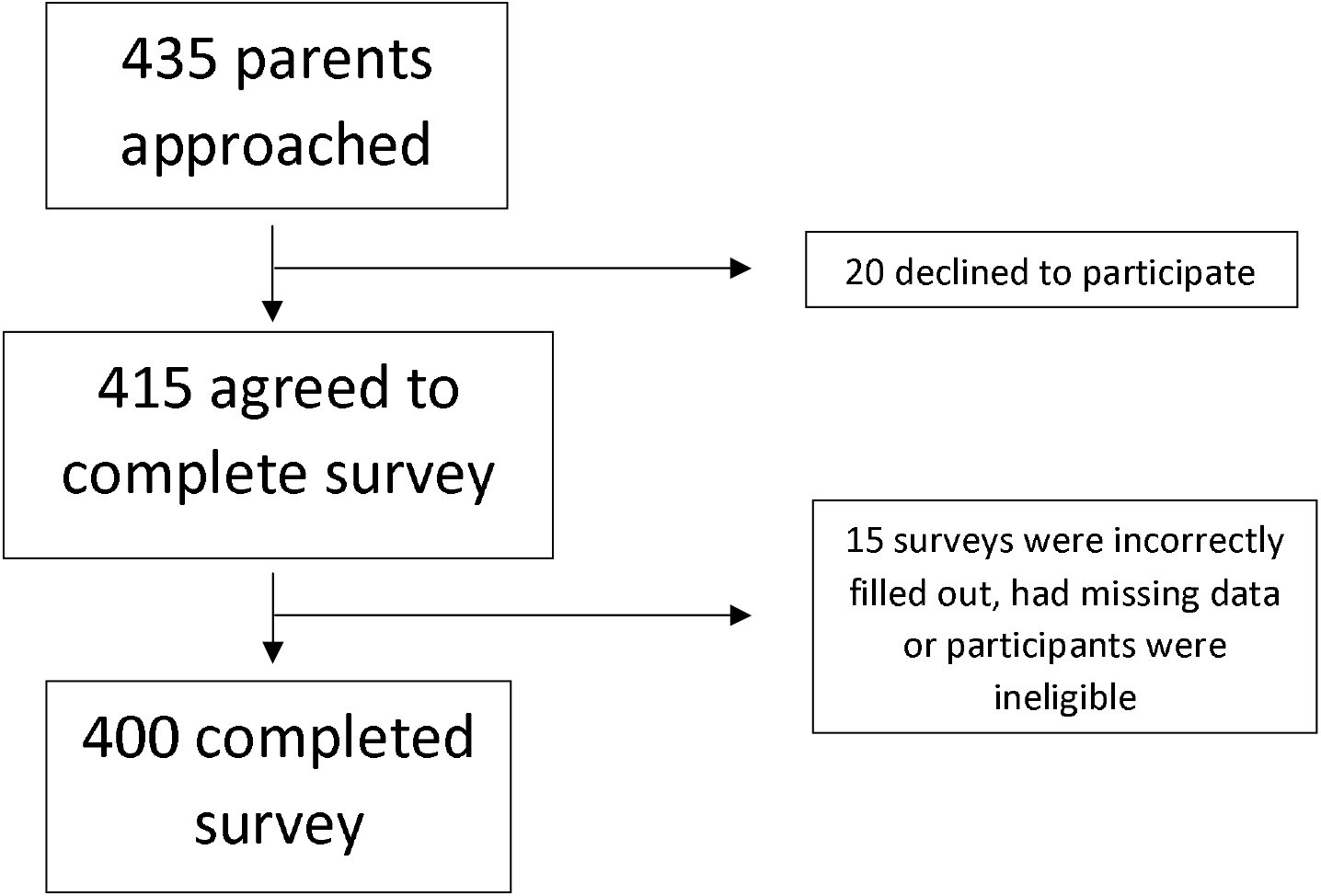

### Patient and public involvement

Patients and the public were not involved in decisions regarding the research question, outcome measures, study design or recruitment to the study. Patients and the public were not asked to assess the burden of the intervention, or time required to participate in the research. However, the survey took approximately 5 minutes to complete and there was no change to the care received, or its duration, for those who were studied. Patients and the public are central to our plans to disseminate the findings of this study.

## Results

### Socio-demographic Data

Overall, there were 400 participants in this study. Of these 224 (56%) were male and 176 (44%) female. The median age was 8 years (range 6 months – 16 years). The mean age was 7.65 years and the modal age was 4 years.

**Figure 1.**
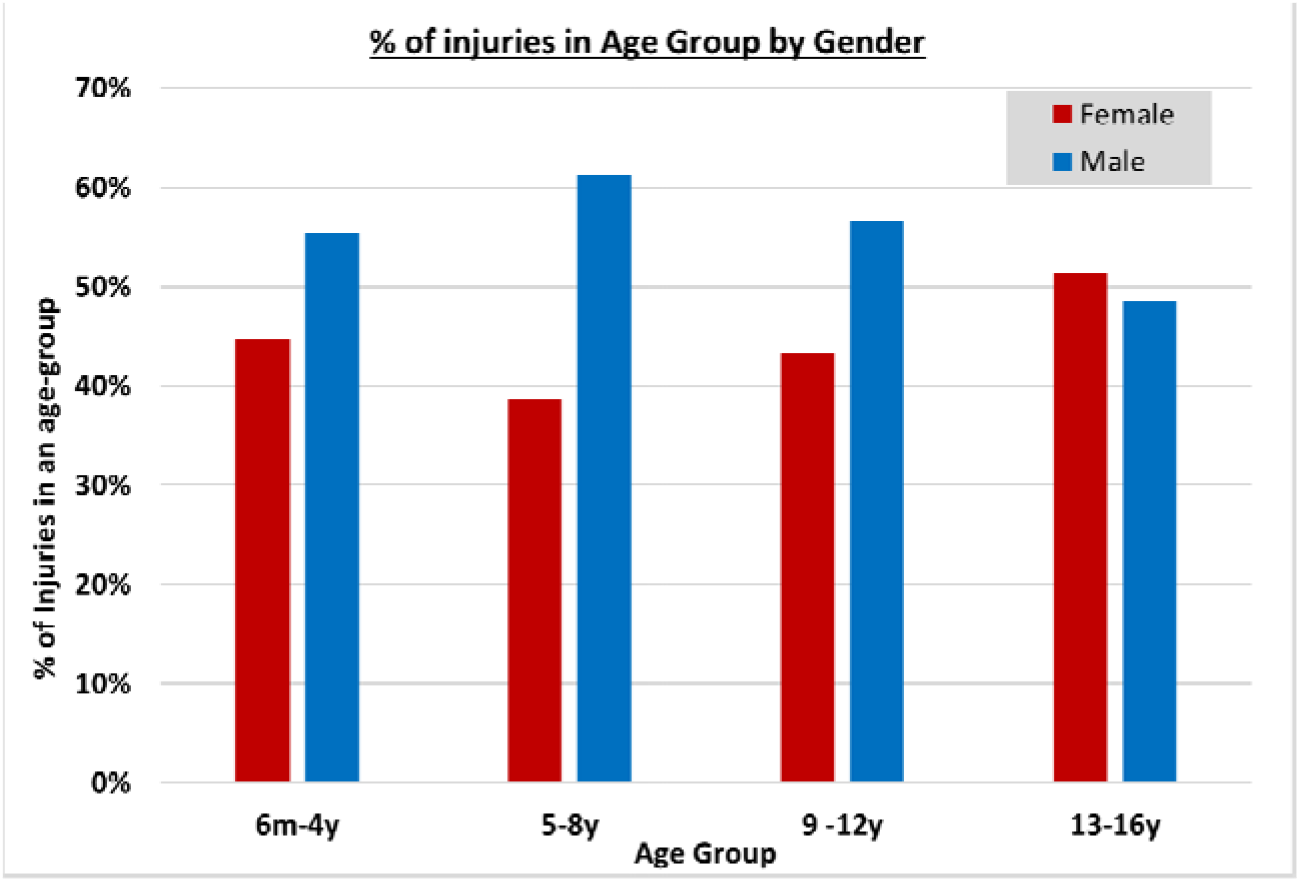

Of the children presenting to ED 332 (83%) had a sibling. Injury was the cause of pain in 259 (64.8%) children and non-injury (e.g. sore throat, headache) was the cause in 141 (35.3%) children.

The most common site of pain was limb in 178 (44.5%) patients, followed by head in 101 (25.3%) patients, abdominal in 81 (20.3%) patients, groin in 16 (4%) patients and chest in 9 (2.3%) patients.

Time from injury/onset of pain until presentation at ED was <48 hours for 341 (85.3%) participants and ≥48 hours for 59 (14.8%) participants.

Pain score was reported using a verbal numerical rating scale (VNRS) from 1-10, 1 being very little pain and 10 being worst pain imaginable. The median pain score was 6. 340 children (85%) reported experiencing moderate to severe pain (i.e. pain score 4).

### Prevalence of Pre-Hospital Analgesia Administration

A total of 189 (47%) children received an analgesic agent prior to arrival at the ED.

### Types and Doses of Analgesia Administered

Of the 400 participants in this study, only 3% did not have any pain-relieving agents at home (i.e. 97% had one or more analgesic agent available). Many households had multiple analgesics, the median number of analgesics per household was 2.18. Paracetamol was the most popular analgesic with 298 (75%) participants having it at home. This was followed by 272 (68%) with ibuprofen and 18 (5%) with solpadol. Overall, 59% of parents administered analgesia where the active compound was paracetamol and 34% administered a form of ibuprofen as analgesia. Liquid suspensions of analgesics are the more popular methods of analgesia administration with 79% parents opting for them over analgesia in tablet form.

**Table 1.** Types and doses of analgesia administered (189/400 patients)

|  |  |  |
| --- | --- | --- |
| **Liquid Paracetamol** |  | **86 (45%)** |
| - 2.5ml | 7 |  |
| - 5ml | 31 |  |
| - 7.5ml | 21 |  |
| - 10ml | 20 |  |
| **Liquid Ibuprofen** |  | **65 (34%)** |
| - 2.5 ml | 4 |  |
| - 5ml | 35 |  |
| - 7.5ml | 18 |  |
| - 10ml | 8 |  |
| **Paracetamol Tablet Form** |  | **26 (14%)** |
| - 250mg | 2 |  |
| - 500mg | 14 |  |
| - 1g | 10 |  |
| **Ibuprofen Tablet Form** |  | **7 (4%)** |
| - 200mg | 3 |  |
| - 400mg | 4 |  |
| **Other** |  | **5 (3%)** |

### Reasons for not Administering Pre-Hospital Analgesia

Altogether 211 (53%) children did not receive any pre-hospital analgesia. The reasons given by parents for not administering pain relief included: 62/211 (29.3%) did not think the child needed it, 39/211 (18.4%) accident did not happen at home, 34/211 (16.1%) did not want to mask the presence of the pain, 20/211 (9.%) believe the hospital should give the medications, 18/211 (8.5%) afraid it would be wrong/harmful. Other reasons included child refused, forgot to, on advice of pharmacist/GP and a preference for natural/homeopathic cures.

Of the parents who did not think their child need pain relief, 75% of these children reported having a moderate-severe pain score (≥4).

**Table 2.** Reasons for withholding analgesia (211/400 patients)

|  |  |  |
| --- | --- | --- |
| **Did not think the child needed it** | **62** | **(29.4%)** |
| Accident did not happen at home | 39 | (18.5%) |
| Did not want to mask the severity of the pain | 34 | (16.1%) |
| Believe the hospital should give the medications | 20 | (9.5%) |
| Afraid it would be wrong/harmful | 18 | (8.5%) |
| On advice of pharmacist/GP | 13 | (6.2%) |
| Other | 25 | (11.8%) |

### Barriers to Pain Management

The following factors were independently associated with an increased likelihood of receiving pre-hospital pain relief: having siblings, a pain score ≥5 and presenting to the emergency department <48 hours from the time of injury/onset of pain.

Of the children without siblings 35.3% (24/68, 95%CI 24%-47%) received pain relief compared to 49.7% (165/332, 95%CI 45%-56%) of children with siblings (x^2^ test, p=0.03, OR 1.811). Children with an estimated pain score of ≤4 received pain relief 29% (30/102, 95%CI 21%-38%) compared to 53.4% (159/298, 95%CI 48%-59%) of children who had a pain score ≥5 (x^2^ test, p=0.004; OR 1.804). Children presenting <48 hours from time of injury/onset of pain received pain relief 50.1% (171/341, 95%CI 45%-55%) cases versus 30.5 % (18/59, 95%CI 19%-42%) children who presented ≥48 hours (x^2^ test, p=0.005, OR 0.436).

**Figure 2.**
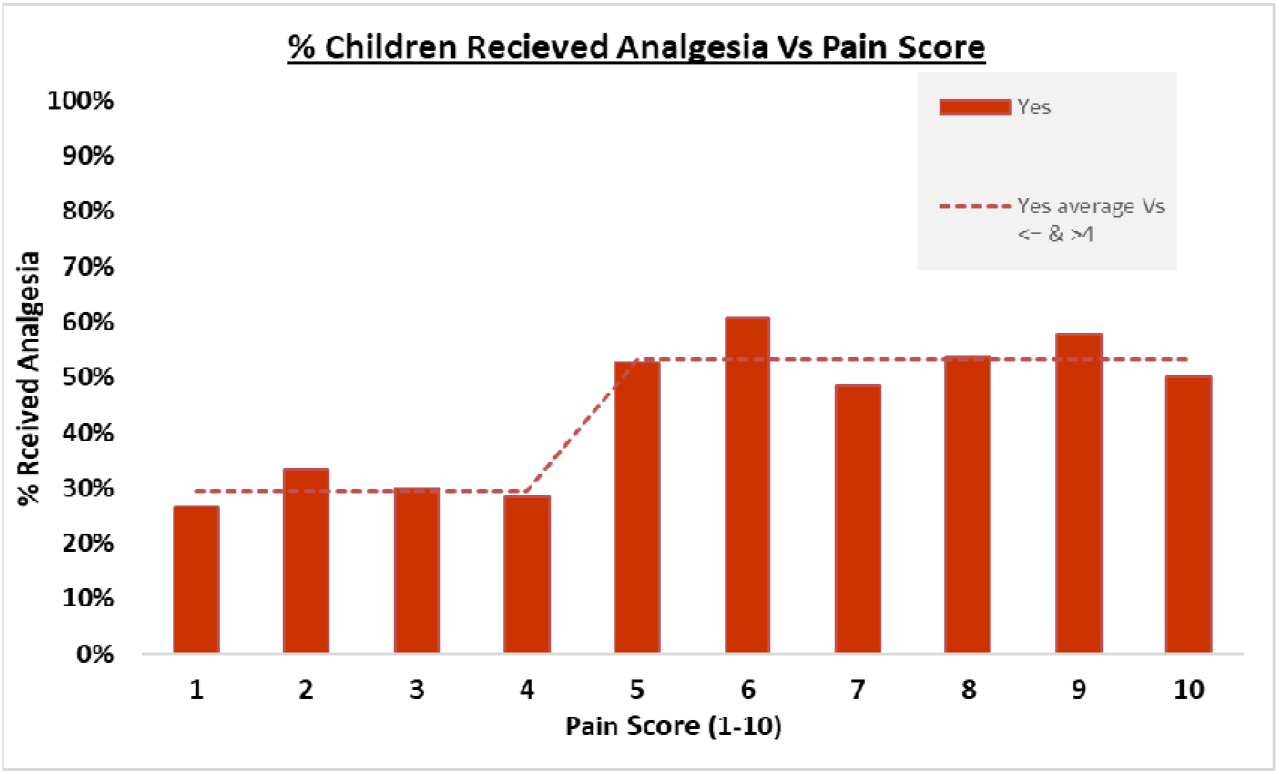

No statistically significant correlation was found between the rate of analgesia administration and sociodemographic factors: age (p=0.804), gender (p=0.114), cause of pain (p=0.079), site of pain (p=0.09 pital m the 7% n %-05, .098).

**Table 3.** Relationship between patient demographics and presenting factors on pre-hospital analgesia administration

|  | Received<br>analgesia | Received<br>no analgesia | OR<br>(95% CI) | % Received analgesia<br>(95% CI) |
| --- | --- | --- | --- | --- |
| **Age** |  |  |  |  |
| 0-4 | 60 | 61 | 0.874 (0.570-1.340) | 50% (41%-58%) |
| 5-8 | 50 | 48 | 0.819 (0.519-1.292) | 51% (41%-61%) |
| 9-12 | 51 | 62 | 1.126 (0.727-1.743) | 45% (36%-54%) |
| 13-16 | 28 | 40 | 1.345 (0.793-2.282) | 41% (29%-53%) |
| **Gender** |  |  |  |  |
| Male | 98 | 126 | 1.152 (0.956-1.374) | 52% (44%-59%) |
| Female | 91 | 85 | 0.837 (0.671-1.044) | 44% (37%-50%) |
| **Siblings** |  |  |  |  |
| Yes | 166 | 166 | 0.907 (0.830-0.990) | 50% (43%-58%) |
| No | 24 | 44 | 1.642 (1.040-2.594) | 35% (29%-42%) |
| **No. of siblings** |  |  |  |  |
| 1 | 70 | 85 | 1.460 (0.946-2.253) | 45% (38%-53%) |
| 2 | 62 | 46 | 0.637 (0.401-1.012) | 57% (48%-67%) |
| 3 | 21 | 25 | 1.216 (0.651-2.271) | 46% (31%-60%) |
| 4 | 9 | 8 | 0.878 (0.330-2.333) | 47% (23%-71%) |
| 5 | 4 | 2 | 0.491 (0.089-2.717) | 33% (-4%-71%) |
| **Time to ED** |  |  |  |  |
| <48 hours | 171 | 170 | 0.890 (0.821-0.965) | 50% (45%-55%) |
| >48 hours | 41 | 18 | 2.040 (1.215-3.426) | 31% (19%-42%) |
| **Cause of Pain** |  |  |  |  |
| Injury | 114 | 145 | 1.139 (0.983-1.320) | 44% (38%-50%) |
| Non-injury | 75 | 66 | 0.788 (0.604-1.029) | 53% (45%-61%) |
| **Site of Pain** |  |  |  |  |
| Limb | 88 | 90 | 0.854 (0.575-1.267) | 49% (42%-57%) |
| Head | 38 | 63 | 1.692 (1.066-2.685) | 38% (28%-47%) |
| Abdominal | 44 | 37 | 0.701 (0.429-1.143) | 54% (43%-65%) |
| Groin | 6 | 10 | 1.517 (0.541-4.258) | 38% (14%-61%) |
| Chest | 3 | 6 | 1.815 (0.447-7.359) | 33% (3%-64%) |
| Other | 10 | 5 | 0.434 (0.146-1.295) | 71% (48%-95%) |
| **Pain Score** |  |  |  |  |
| ≤4 | 30 | 72 | 1.404 (1.109-1.776) | 29% (21%-38%) |
| ≥5 | 159 | 139 | 0.778 (0.656-0.923) | 53% (48%-59%) |

## Discussion

The main finding of this study is that 47% of children presenting to the Emergency Department with acute pain received pre-hospital analgesia. This is an increase compared to the last study of this nature performed in Ireland; Spedding et al, which found just 26% of children received pre-hospital analgesia.^15^ A more recent study by Conrad et al found the rate of pre-hospital analgesia administration to be similar to this study at 54.4%.^16^ This suggests in the 20 years since Spedding et al was published the rates of parental pre-hospital analgesia administration has increased. This is potentially due to parents becoming more educated on the importance of paediatric pre-hospital pain relief.

Overall 53% of children presenting to ED do not receive pre-hospital pain relief and are unnecessarily experiencing oligoanalgesia. Many of the reasons given by parents for not administering analgesia are similar to the barriers identified in previous studies.

The primary reason parents did not administer analgesia was ‘they did not think the child needed it’. However, 75% of these parents reported their child as having a pain score ≥4 (moderate-severe). This indicates there is little correlation between parents believing their child does not require pain relief and their child experiencing a low level of pain. The existing literature has identified how parents frequently under-estimate their child’s pain level and have significant misconceptions regarding how children expressed pain.^17, 18^ A number of other articles have concluded that parent’s consistent underestimation of their child’s pain is a significant barrier to them administering adequate analgesia.^13, 19^

A high proportion of parents reported not administering pain relief due to the belief it would be an inappropriate action i.e. ‘did not want to mask the severity of the pain’ (16.1%), ‘believe the hospital should give the medications’ (9.5%) or ‘afraid it would be wrong or harmful’ (8.5%). Cumulatively 34.1% parents did not administer pain relief due to these fears and misconceptions surrounding the role of pre-hospital analgesia. This finding is supported by previous studies. In Spedding et al 28% parents did not administer analgesia because they were afraid of causing harm.^15^ Maimon et al identified ‘concern that analgesic medication will mask signs and symptoms’ as the primary barrier to parental analgesia administration.^20^ Comparable to this study Conrad et al found 7% parents were ‘afraid it would be wrong or harmful’ and other responses echoed a fear of masking the child’s pain.^16^ Parental misconceptions surrounding analgesic drugs have been identified as significant predictors of inadequate administration of analgesia in the home.^17^ This suggests there is a strong link between parents being misinformed and oligoanalgesia in the pre-hospital setting.

Certain sociodemographic factors are associated with an increased likelihood of parental administration of pre-hospital analgesia. These included: having siblings, presenting to the ED <48 hours and having a pain score ≥5.

This study found a statistically significant association between the child having siblings and parents being more likely to administer analgesia. This may indicate that having more parenting experience increases the likelihood of administering pain relief.

Children who had a pain score ≥5 were more likely to receive pain relief compared to children with a pain score of ≤4. The percentage of children receiving pain relief did not increase linearly with increasing pain score. There was a step-wise increase of analgesia administration between children with a pain score of 4 and 5; identifying this number as the demarcation for parent’s decision to administer analgesia. There is no increase in the probability a child will get pain relieve above a pain score of 5.

Previous studies have found that younger children received analgesia less frequently than older children however no such association was seen in this study.^20, 21^ This finding is supported by Conrad et al which also found no association between age of child and rate of analgesia administration.^16^ In addition no association was found in this study between gender, site of pain or cause of pain and the rate of analgesia administration.

At 97% (389/400) the number of households having one or more analgesic at home was much higher than identified in previous studies. In this study just 3% (11/400) participants had no analgesia at home compared to Spedding et al where 32% parents had no analgesia at home.^15^ Similarly Conrad et al identified having no pain relief at home as a major barrier to analgesia administration, with 12.4% participants selecting it as their reason for withholding pain medication.^16^ In contrast to the existing literature this study demonstrates that where accidents occur at home access to pain relievers is not a barrier to adequate analgesia in most cases.

### Strengths and Limitations

One of the main strengths of this study is its large sample size which increased the statistical power of the results. This study was prospective and not retrospective which eliminated the possibility for documentation bias. All surveys were administered by a single investigator which standardised the process. A pilot study was carried out to ensure the questionnaire was suitable.

This study was limited by a number of factors. It was only conducted across two study sites (EDs of CUH and MUH) so the results may not be generalisable to other locations. Not all eligible participants were recruited to the study. Data collection was limited by the availability of the study investigator, so the sample was not completely randomised; for example, no patients were recruited between 11p.m. and 9a.m. due to the low number of children presenting to ED at night. The survey used had not previously been demonstrated to be reliable or valid.

### Recommendations for Future Practice

Implementation of educational strategies for parents on appropriate paediatric pain management. This should include educating parents on how to appropriately assess their child’s pain, when it is appropriate to administer pain relief and how to select an appropriate type and dose of analgesia for their child. It should also involve informing parents of the potential negative consequences of oligoanalgesia and dispel any misconceptions surrounding analgesics. Organisations which make and sell analgesia for children should be made aware that many parents are underusing their products so they can provide better instruction on how to use them. Any recommendation to advise the public to increase their use of analgesics must be balanced against the risk of abuse or overdose. Educating parents on best practice will empower them to make appropriate decisions based on their individual situation and provide the best care for their child.

## Conclusion

This study has demonstrated that over 50% of children presenting to the ED, with acutely painful conditions, did not receive adequate or timely pain relief pre-ED arrival, causing avoidable suffering. Children were more likely to receive pain relief from parents/guardians if they: have siblings, had a pain score ≥5, presented to ED <48 hours after onset of pain. Parental misconceptions surrounding pain management are a major barrier to them administering pain relief. Further education for parents is required to dispel some of these misconceptions surrounding analgesia and improve the care to this population.

## Data Availability

All de-identified participant data that underlie the results reported in the article will be made available (text, tables, figures and appendices). Study protocol will also be made available. Data will be available beginning immediately following publication with no end date. It will be made available to anyone who wishes to access the data for any purpose. Proposals should be submitted to.

